# Nurses’ Fall Risk Judgements - Cognitive Biases and Contextual Factors Explaining Variability: A multi-centre cross-sectional study

**DOI:** 10.1101/2024.12.03.24318440

**Authors:** Miyuki Takase, Naomi Kisanuki, Sato Yoko, Kazue Mitsunaka, Masako Yamamoto

## Abstract

**Background:** Assessing fall risk is a complex process requiring the integration of diverse information and cognitive strategies. Despite this complexity, few studies have explored how nurses make these judgements. Moreover, existing research suggests variability in nurses’ fall risk assessments, but the reasons for this variation and its appropriateness remain unclear.

**Objective:** This study aimed to investigate how nurses judge fall risk, and the factors associated with their judgements.

**Methods:** Using purposive sampling, 335 nurses from six hospitals in western Japan participated in an online survey. The participants rated the likelihood of falls in 18 patient scenarios and completed measures of base-rate neglect, belief bias, and availability bias. A linear mixed-effects regression tree was used to identify factors related to their judgements, and a linear mixed-effects regression model examined associations between judgement variability, cognitive biases, and clinical specialty.

**Results:** Nurses’ fall risk assessments were primarily influenced by whether patients called for assistance, followed by the use of sleeping pills, the presence of a tube or drain, and patient mobility status. Judgement variability was linked to nurses’ gender, education, clinical specialty, and susceptibility to availability bias.

**Conclusion:** Variability in clinical judgement may be justified when reflecting personalised, context-specific care. However, inconsistencies arising from cognitive biases are problematic. Healthcare organisations should offer targeted training to enhance contextual expertise and reduce the influence of cognitive biases on fall risk assessments.

## Introduction

A patient fall is a major adverse event in healthcare settings. Falls are defined as “inadvertently coming to rest on the ground, floor, or other lower level, excluding intentional changes in position” (World Health Organization, 2007). Each year, a significant number of patients experience falls; for instance, an estimated 700,000 to 1,000,000 falls occur in U.S. hospitals (Agency for Healthcare Research and Quality, 2013), and in Japan, the rate is 2.76 per 1,000 patients (Japan Hospital Association, 2023). Recent studies indicate that 55-60% of patients are at risk of falling (Wu et al., 2019, Yasan et al., 2020), a figure likely to rise as the population ages. Falls can result in severe harm, including death (Takase, 2023), highlighting the importance of addressing this issue. To prevent falls, nurses must assess patients and environmental cues, judge fall risk, and implement appropriate preventive measures (Tanner, 2006).

Nurses’ clinical judgement is crucial in this process, as they are the primary healthcare providers responsible for fall prevention. Clinical judgement involves a reflective and reasoning process, drawing on available data and an extensive knowledge base (Connor et al., 2023). This judgement is a complex cognitive task. Previous studies show that nurses assess fall risk using criteria developed through clinical experience, often relying on cognitive strategies such as adding, subtracting, and weighing various risk factors based on patient characteristics (Takase et al., 2024). Nurses also use fall risk assessment tools, though these have limitations, including variations in criteria and poor predictive validity (Alvarado et al., 2023, Vlaeyen et al., 2021). Assessing fall risk is a complex procedure that requires synthesising information on patient conditions, treatment regimens, and the clinical environment. Despite this complexity, few studies have explored how nurses make these judgements. Moreover, previous research indicates that nurses’ judgements of fall risk can vary (Takase et al., 2024), but it is unclear why this variation occurs or whether it is justified. Such discrepancies can lead to inconsistent care, and if these variations are due to cognitive biases, they may compromise patient safety. Therefore, understanding how nurses make these judgements and identifying the factors that influence them are critical for improving patient care and safety.

## Background

### Nurses’ judgement of risk of falling

The literature identifies various risk factors for falls, which are generally classified into intrinsic (patient-related) and extrinsic (environment-related) categories. Intrinsic factors include sex and age (Wabe et al., 2024), unstable gait and balance (Deandrea et al., 2010, Shao et al., 2023), decline in activities of daily living (Shao et al., 2023), elimination issues such as urgency, frequency of voiding, nocturia, and incontinence (Moon et al., 2021, Noguchi et al., 2016), impaired cognition (Fhon et al., 2023), the use of hypnotics and sedatives (Shao et al., 2023), frailty (Xu et al., 2022), and a tendency to perform tasks independently (Satoh et al., 2022). Extrinsic factors include unsafe footwear (Kobayashi et al., 2017), the presence of tubes and drains (Nakanishi et al., 2021), and wet or slippery floors (Lee et al., 2022).

Nurses must make complex judgements based on available information. For example, they need to evaluate a patient’s fall risk by weighing each risk factor, determining the relative importance of factors when multiple risks are present, and considering how these risks interact when combined. Previous studies have explored the common risk factors nurses perceive (Innab, 2022, Tzeng and Yin, 2013), as well as the alignment between nurses’ assessments of fall risk and patients’ perspectives (Choi et al., 2024). However, no study has investigated the specific criteria or algorithms nurses use to make fall risk judgements. Given that nurses’ assessments directly influence the initiation of preventive interventions (Rice et al., 2022), further research into the methods and decision-making processes they use to judge fall risk is needed.

### Factors related to the variation in judgement

Another issue yet to be fully explored is the source of variation in nursing judgement. Given that clinical judgement involves complex cognitive processes, differences in judgement among healthcare professionals are often inevitable. Variability in clinical judgement and decision-making has been observed among physicians (Hancock et al., 2012, McDonnell et al., 2023, Sutherland and Levesque, 2020), across different professions (Honda et al., 2015, Taggart et al., 2021), between nurses and nursing students (Shinnick and Cabrera-Mino, 2021, Yang and Thompson, 2016), and among nurses themselves (Fernández-de-Maya and Richart-Martínez, 2012, Ferrario, 2003, Thompson et al., 2009). Regarding the fall risk judgement, Vlaeyen et al. (2021) found that the accuracy of risk judgements varied among physiotherapists, nurses, and nurse aides in nursing homes. Similarly, Takase et al. (2024) reported that the fall risk factors deemed critical by nurses differed between individuals.

Several factors may account for these variations, with clinical context playing a significant role. Patient characteristics, presenting symptoms, and treatments differ by clinical setting, influencing which factors nurses prioritise when assessing fall risk. For example, in rehabilitation or aged care wards where many patients have physical limitations, nurses may place greater emphasis on cognitive dysfunction than gait disturbances alone, as patients with cognitive impairments may engage in risky behaviours despite limited mobility. Conversely, in surgical wards, unstable gait might be a higher priority risk due to the presence of post-surgical drains and potential anaemia. Thus, differing clinical contexts necessitate different risk assessments, contributing to variation in nurses’ judgements.

Another potential source of variation in judgement is cognitive bias, defined as systematic tendencies or dispositions that distort information processing, leading to inaccurate judgements or decisions (Crowley et al., 2013, Korteling et al., 2023, Korteling and Toet, 2021). Such biases typically arise when individuals rely on intuition and mental shortcuts (Crowley et al., 2013, Korteling et al., 2023, Korteling and Toet, 2021). While over 50 types of cognitive biases have been identified in healthcare settings (Croskerry and Ryle, 2019), this study focuses on three key biases: base-rate neglect, belief bias, and availability bias. These biases influence probability estimation and logical reasoning, potentially leading to variability in nurses’ judgements.

Base-rate neglect refers to the tendency to ignore underlying incidence rates, prior probabilities, or base rates, either by inflating or reducing them (Croskerry, 2003, Croskerry and Ryle, 2019, Kahneman and Tversky, 1982, O’Sullivan and Schofield, 2018). It is a type of representativeness heuristic (i.e., judging based on how representative A is of B) (Blanco, 2020, Tversky and Kahneman, 1973), and often results from disregarding statistical rules (Ceschi et al., 2019). For example, consider the following.

Among the 1000 people that participated in the study, there were 995 nurses and five doctors. John is a randomly chosen participant in this research. He is 34 years old. He lives in a nice house in a fancy neighbourhood. He expresses himself nicely and is very interested in politics. He invests a lot of time in his career. Which is more likely?

a. John is a nurse.
b. John is a doctor. (Erceg et al., 2022)

Many people chose b) because John’s characteristics seem more representative of a doctor. However, given that 99.5% of participants are nurses, a) is statistically more likely. Similarly, nurses may overlook base rates and overestimate or underestimate specific risks. For instance, a nurse might overestimate the fall risk associated with Medication A after seeing one patient fall while on it, despite the medication rarely causing falls. Since fall risk assessment involves predicting future fall probabilities, adherence to probability principles is essential (Wright et al., 2009, Wright et al., 1994). Ignoring these principles may lead to inaccurate judgements.

Belief bias is “the tendency to accept or reject data depending on one’s personal belief system, especially when the focus is on the conclusion and not the premises or data” (Croskerry and Ryle, 2019). For example, when asked to assess the validity of the following syllogism:

Premise 1: All Eastern countries are communist.

Premise 2: Canada is not an Eastern country.

Conclusion: Canada is not communist. (Erceg et al., 2022)

Many might incorrectly judge this as valid, as the conclusion appears correct despite faulty logic (denying the antecedent). In fall risk assessment, nurses must use sound logic to avoid bias. If they accept seemingly correct conclusions without evaluating the premises, they may misjudge risk. For example, a nurse might think, “A patient with weak legs is likely to fall. Patient A does not have weak legs. Therefore, Patient A will not fall,” which is a logical error.

Availability bias is the tendency to mistakenly judge events as more frequent if they are recent or easily recalled (i.e., readily available in memory) (Croskerry, 2003, O’Sullivan and Schofield, 2018, Tversky and Kahneman, 1973). People tend to recall events that are frequent, recent, distinctive, or associated with other notable events (Tversky and Kahneman, 1973). An example question illustrates this bias:

Which cause of death is more likely?

Commercial airplane crash vs. Bicycle-related (Erceg et al., 2022)

Although the correct answer is “bicycle-related,” many people select “commercial airplane crash” due to its distinctiveness and memorability, especially following recent incidents. Similarly, nurses may overestimate or underestimate risks if certain events come easily to mind. Availability bias is especially relevant in fall risk assessment, as nurses rely on past experiences when judging a patient’s fall risk (Takase et al., 2024). However, basing decisions on memorable but isolated experiences can lead to biased judgements.

While variation due to clinical context may be justifiable, variation due to cognitive biases is not. Thus, identifying and addressing sources of variation is essential. Although a few studies have examined the relationship between clinical context and nurses’ fall risk judgements (e.g., Takase et al., 2024), most studies on cognitive biases in clinical judgement focus on physicians’ diagnostic decisions (e.g., Crowley et al., 2013, Saposnik et al., 2016) or nurses’ assessments of patient acuity or medication administration (Al-Moteri et al., 2022, Essa et al., 2023, Martin et al., 2022). No studies have explored cognitive biases in nurses’ fall risk assessments.

## Aims

This study aimed to address two questions:

1. How do nurses judge the risk of falling?
2. What influences their judgements (i.e., what are the sources of variation in their judgements)?

Answering these questions could inform measures to improve nurses’ fall risk judgements.

## Methods

### Study design

This cross-sectional study is part of a larger investigation into how nurses assess fall risk. The study has two parts: a pilot study (Part 1) and a study addressing the research questions posed above (Part 2). This paper reports the findings of the Part 2 study.

### Participants

Using purposive sampling, participants were recruited from six hospitals (public and private) in western Japan, with capacities ranging from 175 to 740 beds and providing acute, rehabilitation, or extended care. Inclusion criteria required participants to be registered nurses responsible for direct patient care. Exclusion criteria included working in paediatric wards, operating theatres, intensive care units, outpatient departments, or holding managerial roles.

The dependent variable (nurses’ fall risk assessment) consisted of 18 × N (sample size) correlated observations. Regression tree analysis and linear mixed-effects modelling were used to address research questions 1 and 2. For the regression tree analysis, simulation studies indicate that a sample of 1,000 independent observations provides reasonable accuracy and precision (Althnian et al., 2021, Rajput et al., 2023, Sordo and Zeng, 2005). To achieve an effective sample size of 1000 with an intraclass correlation (ICC) of 0.25-0.30, 292-339 participants were needed (see Killip et al., 2004). For the mixed-effects model, sample size simulations using the mixedpower R package (Kumle et al., 2021) indicated that, with b=0.1 for main effects, b=0.05 for interaction terms, and α=0.05, a sample of N=200 would achieve over 80% power. A total of 900 nurses were recruited to meet these requirements.

### Instruments

The following instruments were administered to the participants.

#### Nurses’ fall risk judgements

Patient scenarios were developed to assess nurses’ judgement of fall risk. Each scenario comprised six statements about patient conditions: mobility issues, cognitive impairment and personality, urinary issues, presence of tubes (e.g., intravenous lines and indwelling catheters) or drains, medication use, and age. Each condition had three risk levels (1 = low risk to 3 = high risk) (see Table 1). Scenarios were created following a literature review, consultations with three experienced nurses in risk management, and pilot testing with five academic and three practising nurses. Since the full factorial design would yield 729 scenarios, a fractional orthogonal array was generated using the DoE.base R package (Grömping, 2018), resulting in 18 scenarios (see Supplementary Table 1). Nurses rated fall probability from 0-100% in 5% increments, which were later converted to a probability of 0-1.00 in 0.05 increments for the analysis.

**Table 1.** The structure of patient scenarios and examples.

| Risk level | Conditions |  |  |  |  |  |
| --- | --- | --- | --- | --- | --- | --- |
|  | Mobility issues | Cognitive impairment and personality (Does not call nurses) | Urinary issues | Presence of tubes or drains | Medication use | Age |
| 1 | Independent | No cognitive impairment and calls nurses when necessary | No urinary problems | No tube or drain inserted | Antibiotic | Less than 65 years old |
| 2 | Walks alone but unstable | No cognitive impairment, but reserved and does not call nurses when necessary | Nocturia | One tube or drain inserted | Anti-hypertensive | 65-79 years old |
| 3 | Needs assistance in transfer | Impaired cognition and does not call nurses when necessary | Urinary frequency | Multiple tubes and/or drains inserted | Sleeping pill | 80 years old and over |

Example: Please read the following 18 scenarios and assess on a scale of 0-100% the probability that the patient shown in each scenario will fall. Note that all cases are male patients with no history of falls.
1) Mr. A is a patient who exhibits the following characteristics<sup>#</sup>.
- (1) Urinates at night. - (2) Takes a sleeping pill. - (3) Under 65 years old. - (4) Needs assistance in transfer. - (5) Has no drain or tube inserted. - (6) Has no cognitive impairment, but reserved and does not call nurses when necessary.
Note: <sup>#</sup> In the scenarios, patients' characteristics are presented in a random order.

#### Cognitive biases

Existing scales for cognitive biases contain general statements, with some items irrelevant in Japanese settings. Therefore, new scales were developed specifically for fall risk judgements, targeting base-rate neglect, belief bias, and availability biases, based on established scales. Each scale had three items with two response choices—one indicating bias (scored 1) and the other non-bias (scored 0). The scales were pilot-tested with five academics and three nurses, and then administered to 55 nurses for reliability testing. The Kuder-Richardson 20 (KR20) coefficients of base-rate neglect, belief and availability biases were 0.70, 0.65, and 0.41 respectively. The reliability of the availability bias was low, but an item response analysis showed good discriminant validity. Moreover, a reliable measure of availability bias is lacking (Berthet, 2021), especially in the Japanese context. Thus, we retained the original items. In all the cognitive bias scales, responses were aggregated for a total score from 0 to 3; scores of 2 or more indicated cognitive bias. Sample questions are presented in Table 2.

**Table 2.** Sample items of the cognitive bias scale.

|  |
| --- |
| <u>Base-rate neglect</u><br>Hospital B has 1,000 patient falls each year. About 50 of these falls are due to vertigo, and the remaining 950 are due to unstable gait. Ms. Yasuda is a randomly selected patient at Hospital B. She is a 90-year-old woman admitted for treatment of heart failure. Her face is pale, and when she stands up, she complains, “Sometimes my eyes go completely black”. She sometimes wobbles. Which is more likely?<br>1. Ms. Yasuda falls because of vertigo. (Biased response)<br>2. Ms. Yasuda falls because of unstable gait. (Correct response) |
| <u>Belief bias</u><br>We will now solve syllogisms. First, two premises are presented, following a conclusion. You are to determine if the conclusion follows logically from the premises. Assume that all of the premises are true. Now read the following syllogism and choose 1 if it is logically correct or 2 if it is incorrect.<br>Premise 1: Patients who can move on their own are prone to falls.<br>Premise 2: Ms. A cannot move by herself and is bedridden.<br>Conclusion: Therefore, her risk of falling is low.<br>1. Correct (Biased response)<br>2. Incorrect (Correct response) |
| <u>Availability bias</u><br>Which do you think leads to more deaths? Please choose the one you think causes more deaths.<br>1. Fall (Correct response)<br>2. Traffic accident (Biased response) |

#### Demographics

These questions gathered data on participants’ age, sex, highest educational degree, years of clinical experience, and current specialty.

#### Data collection procedures

After receiving approval from each institution’s Director of Nursing, invitation letters were distributed to potential participants in August, 2024. The letter explained the study’s purpose, methods, and ethical considerations, and provided a QR code and a URL link to a Google Forms survey. Interested participants were directed to the online survey site to complete the questionnaire, with consent assumed upon submission. Participants had four weeks to complete the questionnaire.

#### Analysis

Exploratory factor analysis based on a tetrachoric correlation matrix with oblique rotation and KR20 were computed to assess the construct validity and internal consistency of the cognitive bias scales. Descriptive statistics were then computed, and variable distributions were checked. As will be shown, there was a small portion of missing responses in some question items (2.4% at maximum). Thus, the subsequent analyses were conducted by replacing missing values, i.e., those in the continuous or ordinal variables were replaced with the median values, while those in categorical variables were replaced with the most representative category.

To explore how nurses assess fall risk (research question 1), a regression tree analysis was performed. Each nurse provided 18 responses, one for each patient scenario, resulting in correlated data. Thus, a linear mixed-effects model tree with a random intercept was fitted to account for intraclass correlation, followed by 10-fold cross-validation to calculate explained variance. The glmertree R package (Fokkema et al., 2018) was used.

To identify sources of variation in nurses’ judgements (research question 2), a mixed-effects regression model was applied. This model included dichotomised responses on cognitive biases, risk variables (Table 1), clinical specialty, and demographic factors as main effects, along with interaction terms between risk variables and clinical specialty to examine if weighting differed by specialty. Risk variables were collapsed into two levels (combining levels 2 and 3) and clinical specialties into six categories to reduce comparisons and avoid overfitting. Analyses were performed using StataNow (version 18.0, StataCorp, Texas, USA) and R (version 4.4.1).

#### Ethical considerations

This study complies with the Declaration of Helsinki developed by the World Medical Association, and approval was obtained from the institutional review board prior to the data collection.

## Results

A total of 337 nurses completed the online survey, yielding a response rate of 37.4%. Of these, two nurses did not provide their judgements on the patient scenarios, leaving 335 questionnaires for further analysis. The characteristics of the participants are described in Table 3.

**Table 3.** Characteristics of the participants.

| Variables |  | Mean/Frequency | Standard deviation/Proportion |
| --- | --- | --- | --- |
| Age |  | 34.06 | 10.97 |
| Length of experience |  | 10.52 | 9.45 |
| Sex | Male | 35 | 9.86% |
|  | Female | 298 | 88.96% |
|  | Missing | 2 | 0.60% |
| Education | Diploma | 212 | 63.28% |
|  | Bachelor & Postgraduate | 119 | 35.52% |
|  | Missing | 4 | 1.19% |
| Clinical specialty<sup>#</sup> | Respiratory/Cardiovascular wards | 28 | 8.36% |
|  | Convalescent rehabilitation/Community-based Care wards | 87 | 25.98% |
|  | Orthopaedic /Neurology wards | 30 | 8.96% |
|  | Long-term/Palliative care wards | 46 | 13.74% |
|  | Gastrointestinal wards | 45 | 13.44% |
|  | Others | 93 | 27.77% |
|  | Missing | 6 | 1.8% |

Note: There are six missing values (1.8%) in age and eight missing values (2.4%) in the length of experience.
<sup>#</sup> Nurses were categorised into six groups based on their main clinical specialty.

Figure 1 presents the nurses’ judgements on the risk of falling, represented as the estimated probability of falling on a scale from 0 to 1. Scenario 8 (i.e., a patient with a risk level of 3 across all variables) was rated as the highest risk, while Scenario 17 (i.e., a patient with a risk level of 1 across all variables) was rated as the lowest. The ICC of judgements within nurses was 0.279 (standard error = 0.018).

**Figure 1.**
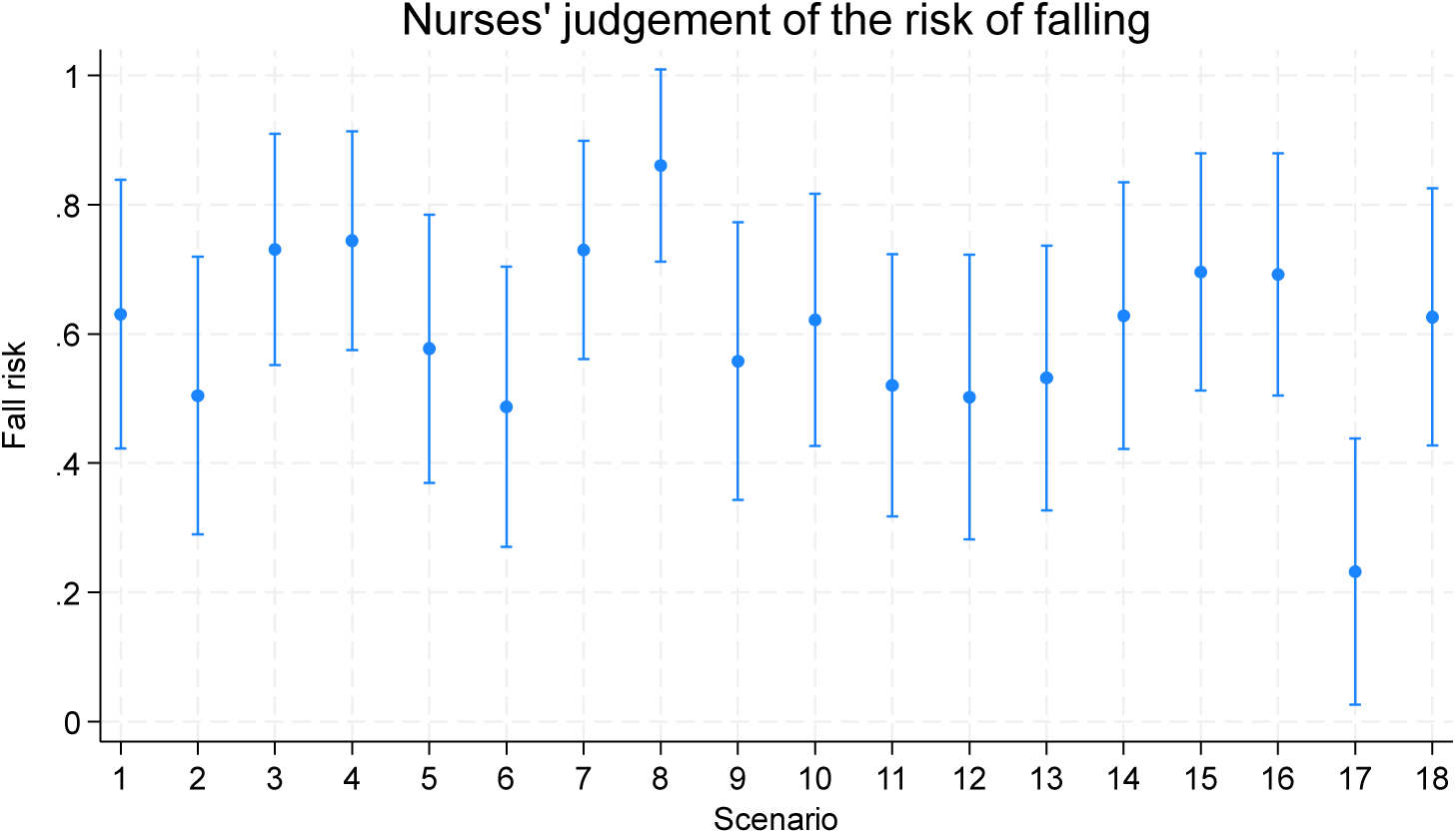
The means and standard deviation of nurses’ judgement of the risk of falling in each scenario

Supplementary Table 2 presents the results of factor analysis on the cognitive bias scales, along with their reliability coefficients. As shown in the table, three factors representing base-rate neglect, belief bias, and availability bias were identified, explaining 64.3% of the variance. However, the KR20 coefficients were low, particularly for the base-rate neglect (KR20 = 0.323) and the belief bias scale (KR20 = 0.434). These low reliabilities were due to the majority of nurses consistently choosing biased responses for base-rate neglect (68.66%) and belief bias items (35.52%), resulting in small variances (see Supplementary Table 3). An opposite trend was observed in the availability bias scale.

Figure 2 presents the results of a regression tree analysis, illustrating how nurses judged the risk of patient falls. The first variable used to differentiate between the nurses’ judgements was the patient’s inability to call for assistance due to cognitive impairment. If a patient could not call for help, the second variable used to differentiate between the judgements was whether the patient had taken a sleeping pill, followed by whether the patient had stable or unstable gait. When a patient had cognitive impairment, could not call for assistance, was taking a sleeping pill, and did not have an unstable gait (but required assistance with ambulation), nurses’ judgement of fall risk was highest (see Node 28). Scenario 8 represents this patient’s characteristics.

**Figure 2.**
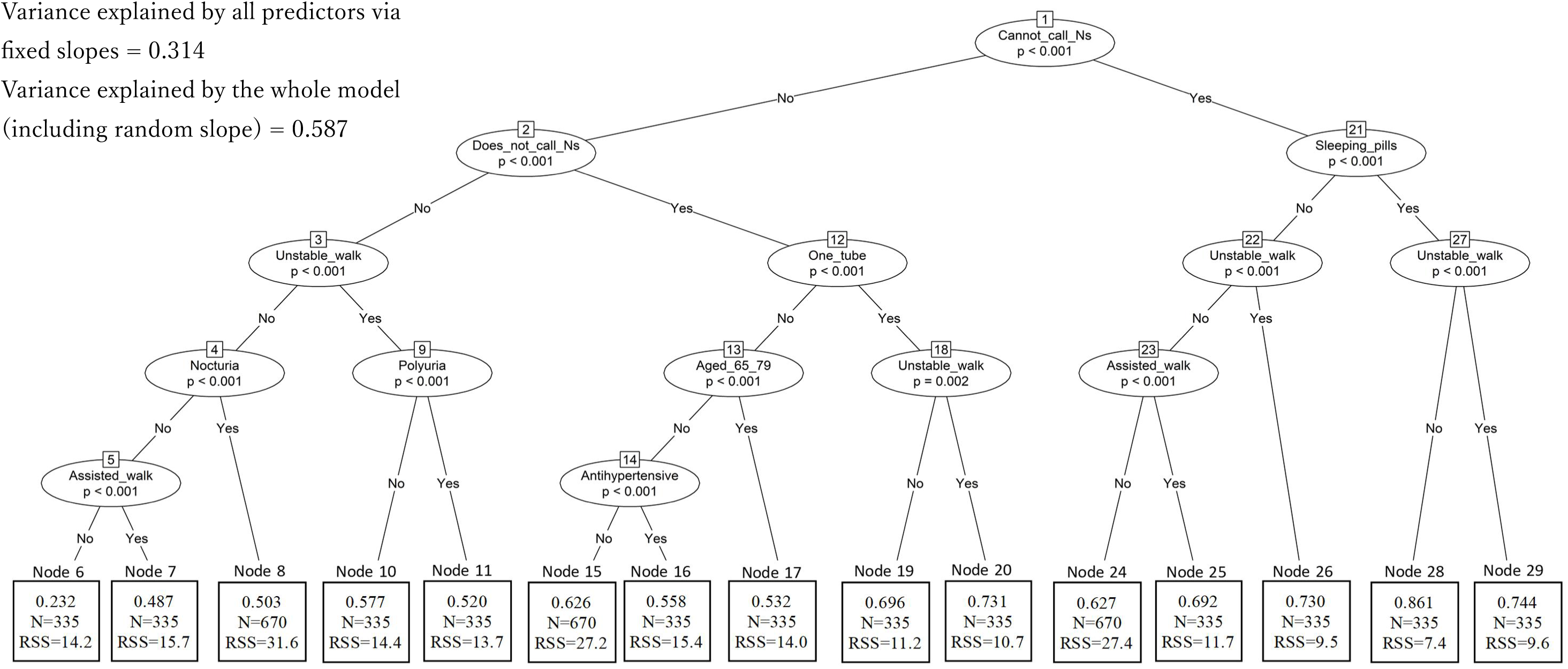
Regression tree illustrating nurses’ judgement in the risk of falling Note: The numeric values in the boxes indicate the mean probability of falling estimated by nurses, and ‘RSS’ indicates the residual sum of squares. N indicates the number of observations (335×18=6030 observations in total). In this analysis, patient conditions (see Table 1) were dummy-coded, with the risk level 1 variable serving as the reference category for each condition.

Nurses judged the risk of falling as the second highest when a patient had cognitive impairment, took a sleeping pill, and had an unstable gait (see Node 29, representing Scenario 4). The next highest judgements of fall risk were for a patient with cognitive impairment and unstable gait (Node 26, representing Scenario 7), and for a patient who was hesitant to call nurses, had one tube/drain, and an unstable gait (Node 20, representing Scenario 3).

The regression tree model showed that nurses assigned progressively lower probabilities of falls to the following risk factors in descending order: patients’ age, intake of anti-hypertensive agents, and presence of urinary issues. Only the need for assistance with transfers was associated with a fall probability below 50% (see Node 7), while a patient without any of these conditions was still judged to have a small chance of falling (23.2%).

Finally, the regression tree model suggests that there was variation in nurses’ judgement. In fact, the tree accounts for 58.7% of the variance, while a large portion (27.3%) was explained by a random effect.

A mixed-effects regression model was used to identify factors that could be associated with this variation (see Table 4; see also Supplementary Table 4 for a correlation analysis). The results indicated that participants’ sex, educational background, clinical specialty, and susceptibility to availability bias were associated with differences in nurses’ judgement of fall risk, along with the presence of the risk factors listed in Table 1. Female nurses and those with a diploma tended to judge the risk of falling to be higher than their counterparts (6.7% and 4.7% respectively). Nurses working in convalescent rehabilitation or community-based care wards judged patients’ risk 9.3% to be higher than those in other specialties. Additionally, nurses more susceptible to availability bias tended to judge the risk as 3% lower than those who were less susceptible.

**Table 4.**
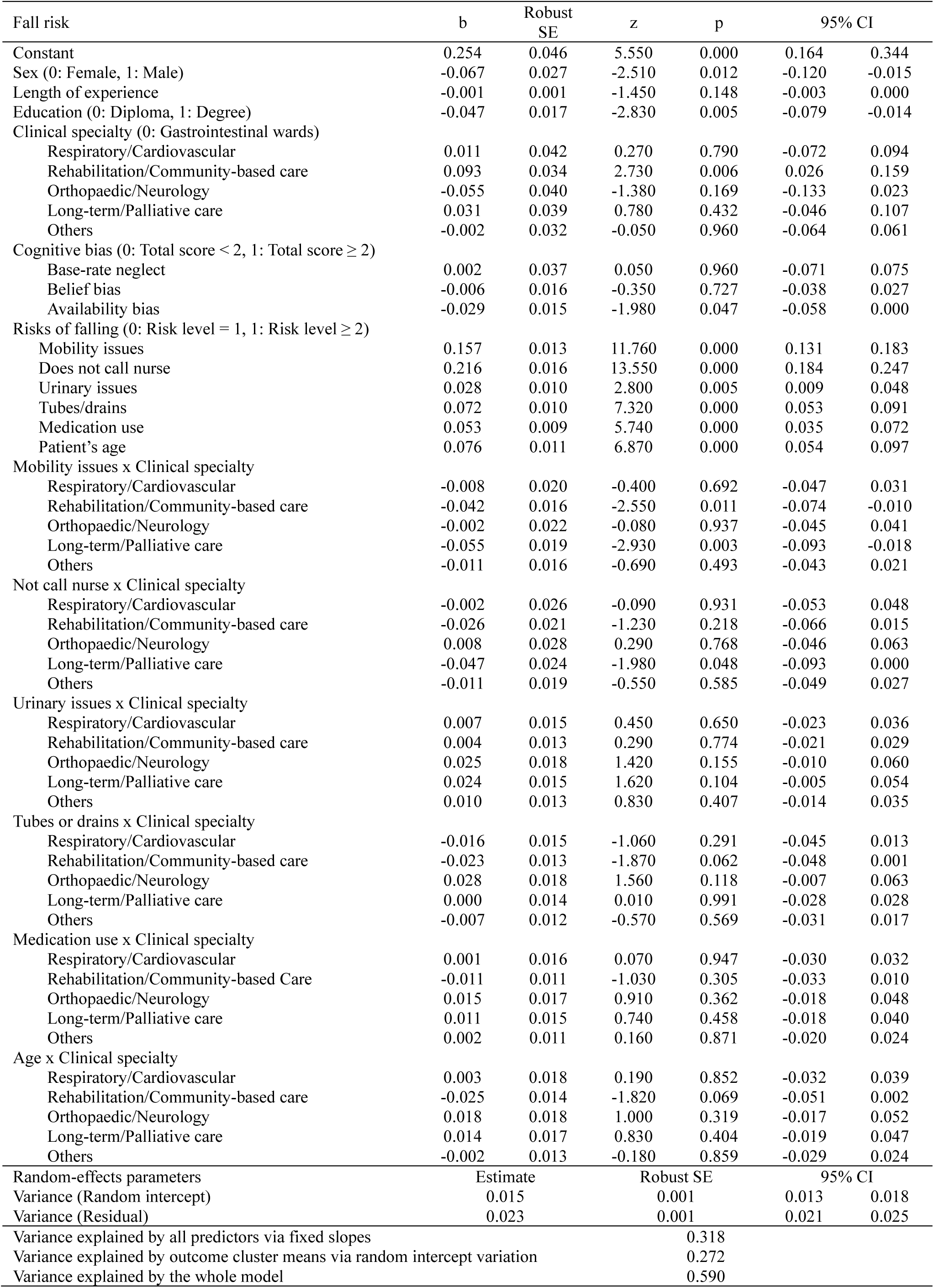
The results of a mixed-effect model.

| Fall risk | b | Robust SE | z | p | 95% CI |  |
| --- | --- | --- | --- | --- | --- | --- |
| Constant | 0.254 | 0.046 | 5.550 | 0.000 | 0.164 | 0.344 |
| Sex (0: Female, 1: Male) | -0.067 | 0.027 | -2.510 | 0.012 | -0.120 | -0.015 |
| Length of experience | -0.001 | 0.001 | -1.450 | 0.148 | -0.003 | 0.000 |
| Education (0: Diploma, 1: Degree) | -0.047 | 0.017 | -2.830 | 0.005 | -0.079 | -0.014 |
| Clinical specialty (0: Gastrointestinal wards) |  |  |  |  |  |  |
| Respiratory/Cardiovascular | 0.011 | 0.042 | 0.270 | 0.790 | -0.072 | 0.094 |
| Rehabilitation/Community-based care | 0.093 | 0.034 | 2.730 | 0.006 | 0.026 | 0.159 |
| Orthopaedic/Neurology | -0.055 | 0.040 | -1.380 | 0.169 | -0.133 | 0.023 |
| Long-term/Palliative care | 0.031 | 0.039 | 0.780 | 0.432 | -0.046 | 0.107 |
| Others | -0.002 | 0.032 | -0.050 | 0.960 | -0.064 | 0.061 |
| Cognitive bias (0: Total score < 2, 1: Total score ≥ 2) |  |  |  |  |  |  |
| Base-rate neglect | 0.002 | 0.037 | 0.050 | 0.960 | -0.071 | 0.075 |
| Belief bias | -0.006 | 0.016 | -0.350 | 0.727 | -0.038 | 0.027 |
| Availability bias | -0.029 | 0.015 | -1.980 | 0.047 | -0.058 | 0.000 |
| Risks of falling (0: Risk level = 1, 1: Risk level ≥ 2) |  |  |  |  |  |  |
| Mobility issues | 0.157 | 0.013 | 11.760 | 0.000 | 0.131 | 0.183 |
| Does not call nurse | 0.216 | 0.016 | 13.550 | 0.000 | 0.184 | 0.247 |
| Urinary issues | 0.028 | 0.010 | 2.800 | 0.005 | 0.009 | 0.048 |
| Tubes/drains | 0.072 | 0.010 | 7.320 | 0.000 | 0.053 | 0.091 |
| Medication use | 0.053 | 0.009 | 5.740 | 0.000 | 0.035 | 0.072 |
| Patient's age | 0.076 | 0.011 | 6.870 | 0.000 | 0.054 | 0.097 |
| Mobility issues x Clinical specialty |  |  |  |  |  |  |
| Respiratory/Cardiovascular | -0.008 | 0.020 | -0.400 | 0.692 | -0.047 | 0.031 |
| Rehabilitation/Community-based care | -0.042 | 0.016 | -2.550 | 0.011 | -0.074 | -0.010 |
| Orthopaedic/Neurology | -0.002 | 0.022 | -0.080 | 0.937 | -0.045 | 0.041 |
| Long-term/Palliative care | -0.055 | 0.019 | -2.930 | 0.003 | -0.093 | -0.018 |
| Others | -0.011 | 0.016 | -0.690 | 0.493 | -0.043 | 0.021 |
| Not call nurse x Clinical specialty |  |  |  |  |  |  |
| Respiratory/Cardiovascular | -0.002 | 0.026 | -0.090 | 0.931 | -0.053 | 0.048 |
| Rehabilitation/Community-based care | -0.026 | 0.021 | -1.230 | 0.218 | -0.066 | 0.015 |
| Orthopaedic/Neurology | 0.008 | 0.028 | 0.290 | 0.768 | -0.046 | 0.063 |
| Long-term/Palliative care | -0.047 | 0.024 | -1.980 | 0.048 | -0.093 | 0.000 |
| Others | -0.011 | 0.019 | -0.550 | 0.585 | -0.049 | 0.027 |
| Urinary issues x Clinical specialty |  |  |  |  |  |  |
| Respiratory/Cardiovascular | 0.007 | 0.015 | 0.450 | 0.650 | -0.023 | 0.036 |
| Rehabilitation/Community-based care | 0.004 | 0.013 | 0.290 | 0.774 | -0.021 | 0.029 |
| Orthopaedic/Neurology | 0.025 | 0.018 | 1.420 | 0.155 | -0.010 | 0.060 |
| Long-term/Palliative care | 0.024 | 0.015 | 1.620 | 0.104 | -0.005 | 0.054 |
| Others | 0.010 | 0.013 | 0.830 | 0.407 | -0.014 | 0.035 |
| Tubes or drains x Clinical specialty |  |  |  |  |  |  |
| Respiratory/Cardiovascular | -0.016 | 0.015 | -1.060 | 0.291 | -0.045 | 0.013 |
| Rehabilitation/Community-based care | -0.023 | 0.013 | -1.870 | 0.062 | -0.048 | 0.001 |
| Orthopaedic/Neurology | 0.028 | 0.018 | 1.560 | 0.118 | -0.007 | 0.063 |
| Long-term/Palliative care | 0.000 | 0.014 | 0.010 | 0.991 | -0.028 | 0.028 |
| Others | -0.007 | 0.012 | -0.570 | 0.569 | -0.031 | 0.017 |
| Medication use x Clinical specialty |  |  |  |  |  |  |
| Respiratory/Cardiovascular | 0.001 | 0.016 | 0.070 | 0.947 | -0.030 | 0.032 |
| Rehabilitation/Community-based Care | -0.011 | 0.011 | -1.030 | 0.305 | -0.033 | 0.010 |
| Orthopaedic/Neurology | 0.015 | 0.017 | 0.910 | 0.362 | -0.018 | 0.048 |
| Long-term/Palliative care | 0.011 | 0.015 | 0.740 | 0.458 | -0.018 | 0.040 |
| Others | 0.002 | 0.011 | 0.160 | 0.871 | -0.020 | 0.024 |
| Age x Clinical specialty |  |  |  |  |  |  |
| Respiratory/Cardiovascular | 0.003 | 0.018 | 0.190 | 0.852 | -0.032 | 0.039 |
| Rehabilitation/Community-based care | -0.025 | 0.014 | -1.820 | 0.069 | -0.051 | 0.002 |
| Orthopaedic/Neurology | 0.018 | 0.018 | 1.000 | 0.319 | -0.017 | 0.052 |
| Long-term/Palliative care | 0.014 | 0.017 | 0.830 | 0.404 | -0.019 | 0.047 |
| Others | -0.002 | 0.013 | -0.180 | 0.859 | -0.029 | 0.024 |
| Random-effects parameters | Estimate |  | Robust SE |  | 95% CI |  |
| Variance (Random intercept) | 0.015 |  | 0.001 |  | 0.013 | 0.018 |
| Variance (Residual) | 0.023 |  | 0.001 |  | 0.021 | 0.025 |
| Variance explained by all predictors via fixed slopes |  |  |  |  | 0.318 |  |
| Variance explained by outcome cluster means via random intercept variation |  |  |  |  | 0.272 |  |
| Variance explained by the whole model |  |  |  |  | 0.590 |  |

Several interaction terms were also significant. Compared to nurses working in gastrointestinal wards, the judgements of nurses in convalescent rehabilitation, community-based care, long-term care, and palliative care wards were less associated with patients’ mobility issues (see Figures 3a and 3b). The judgements of nurses specialising in long-term and palliative care were also less associated with whether patients called for assistance, compared to those in gastrointestinal wards (see Figure 3c). Although not reaching p<0.05, rehabilitation and community-based care nurses showed a tendency to attach less importance to whether patients had tubes/drains and were over 65 years old, when judging the risk of falling.

**Figure 3.**
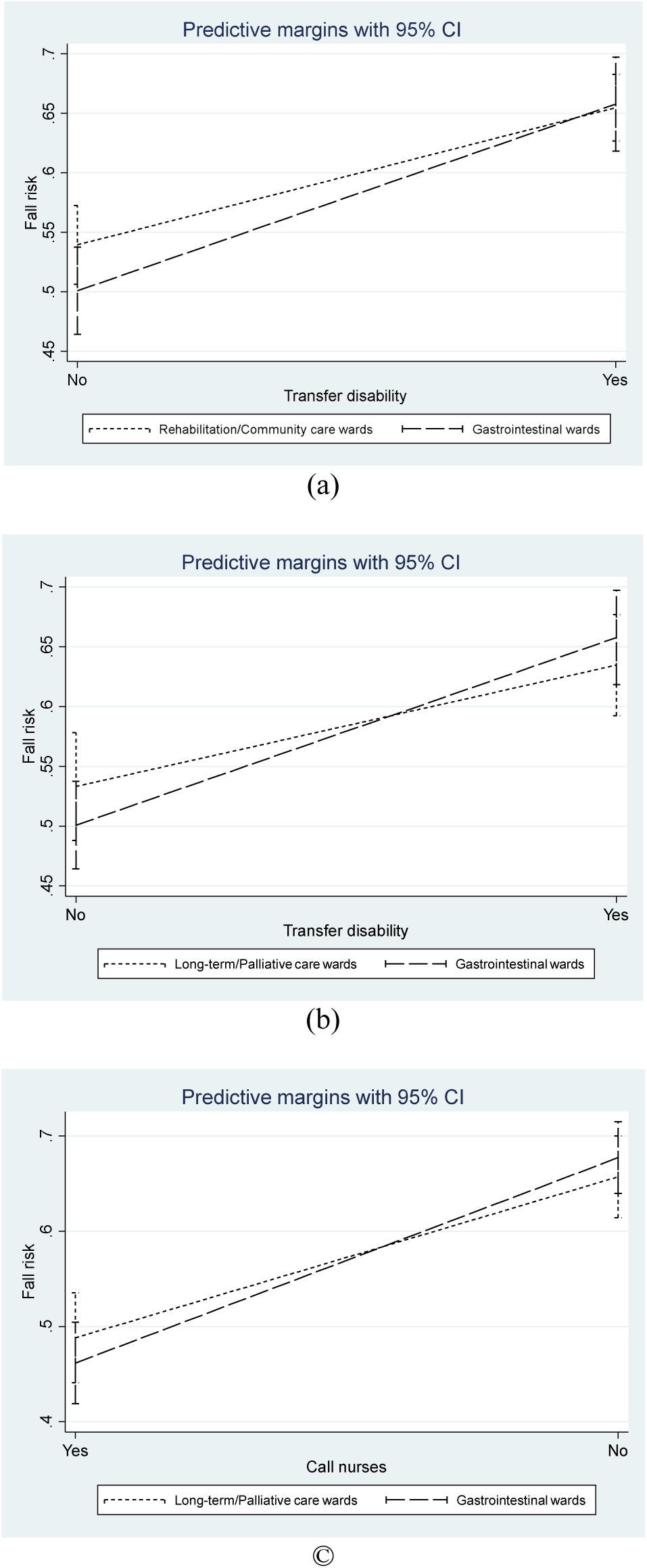
Moderating effects between the risk factors and clinical specialty

## Discussion

This study examined how nurses judge fall risk, with attention to variations related to their clinical specialty and tendencies toward cognitive biases. Regression tree analysis revealed that nurses’ judgements were primarily based on whether patients called for assistance when needed, followed by other fall-related factors such as the use of sleeping pills, mobility disorders, the presence of medical devices, and urinary issues. Patients with cognitive impairment, inability to call for help, use of sleeping pills, and mobility disorders were identified as being at the highest risk. Previous studies have shown that most falls are unwitnessed (Francis-Coad et al., 2020, Takase, 2023), and tend to happen to patients inclined to act independently (Satoh et al., 2023). Additionally, factors such as hypnotic use, cognitive impairment, and mobility disorders have been consistently associated with fall risk in numerous studies, including a recent systematic review (Heinzmann et al., 2024). Consequently, patients who engage in activities independently, despite underlying fall risks, are perceived as being particularly vulnerable to falling. Nurses’ belief that falls can be prevented by accompanying patients highlights their emphasis on a patient’s ability to seek nursing assistance as a primary consideration in fall risk assessment.

Patients who did not call for assistance despite lacking cognitive impairment, having a tube or drain, and experiencing gait issues (e.g., unstable walking or requiring transfer assistance) were also judged by nurses to have a high risk of falling. One study reported that approximately 90% of patients at risk of falling had a tube or drain inserted (Sakai et al., 2016). Given that most participants in this study worked in acute care hospitals, where many patients have tubes or drains, nurses’ heightened concern about the impact of these devices on fall risk (Takase et al., 2024) is understandable.

While mobility issues were identified as a key factor in nurses’ fall risk judgements, requiring assistance with transfers alone was associated with less than a 50% perceived risk of falling. Several explanations may account for this. First, studies suggest that fall rates do not always correspond to fall frequency (Inoue et al., 2024). Thus, even if mobility impairment is recognised as a high-risk factor, the frequency of falls may be low if few patients exhibit such impairments. Given that the majority worked in acute settings, the number of patients requiring transfer assistance may have been small, leading to fewer incidents. This could create an impression among nurses that fall risk is moderate. Second, nurses may feel confident that falls can be prevented if patients call for assistance. Finally, mobility issues may be viewed as amplifying rather than independently driving fall risk. For instance, a study found that the presence of mobility impairments heightened fall risk in cognitively impaired patients (Martin et al., 2013). Consequently, in the absence of other risk factors, the overall risk may be considered moderate.

The results also revealed variation in nurses’ judgements, which usually stemmed from differences in how they weighed each risk factor (Stamp, 2012, Thompson et al., 2009). In this study, factors such as nurses’ gender, educational background, clinical specialty, and susceptibility to availability bias were associated with how they evaluated each risk. The mixed-effects regression model indicated that nurses in convalescent rehabilitation and community-based care wards rated patients’ fall risk significantly higher than those on gastrointestinal wards. In these settings, patients are in the process of regaining mobility and transitioning back to their communities, which inherently increases their fall risk (Kato et al., 2022). High fall rates in convalescent rehabilitation wards have been reported in other studies as well (Ainuddin et al., 2019, Kato et al., 2022). By contextualising each scenario within their practice, nurses in these wards might have assessed fall risk more severely than their gastrointestinal counterparts.

Additionally, nurses in convalescent rehabilitation/community-based care, and long-term/palliative care wards perceived the risks associated with mobility disorders and the inability to call for assistance differently from nurses in gastrointestinal settings. For nurses in rehabilitation and community care, mobility disorders had less impact on their judgement compared to those in gastrointestinal nursing. This could be explained by two factors. First, nurses in rehabilitation and community care settings may perceive mobility limitations as universal among their patients, thus not significantly influencing fall risk. A recent study found no difference in mobility between patients who fell and those who did not in stroke rehabilitation (Hong et al., 2024). Alternatively, nurses may believe that patients in these settings have an inherent fall risk, regardless of their mobility status. Kato et al. (2022) found that although patients with functional impairments had a higher risk of falling, recovery from mobility impairment led to greater activity levels, which resulted in increased fall rates over time. This may explain why nurses rated patients without mobility issues as higher risk than their gastrointestinal counterparts.

Similarly, nurses in long-term/palliative care rated the impact on fall risk of mobility disorders and the inability to call for assistance lower than nurses in gastrointestinal nursing. This may be because nurses in long-term/palliative care settings may not view cognitive or mobility impairments as distinguishing factors for fall risk, as the majority of patients in these wards experience these conditions. Additionally, patients in long-term/palliative care are often frail, dependent, depressed, and suffer from multiple chronic comorbidities, such as congestive heart failure and chronic pulmonary diseases, all of which are associated with increased fall risk (Oren et al., 2022, Prabhakaran et al., 2020, Xu et al., 2022). Thus, a single risk factor may not carry as much weight as it does in other specialties. In contrast, on acute care wards like gastrointestinal medicine and surgery, diseases and treatments can dramatically alter patients’ mobility and cognitive function (Milisen et al., 2012, Perell et al., 2001). Consequently, changes in functional status have more visible impacts on fall risk. These differing characteristics of clinical specialties probably explain the variation in how these risk factors are prioritised.

While variation due to contextual clinical judgement may be justified, variation stemming from individual factors such as gender, educational background, and susceptibility to cognitive biases is not justifiable. This is because there is no clinical evidence to suggest that such factors affect fall risk. Gender differences in risk perception are well-documented, with studies consistently showing that females tend to perceive greater susceptibility to and severity of various risk factors (Brown et al., 2021, Harris and Jenkins, 2006, Kim et al., 2018), often resulting in them recommending more preventive measures compared to their male counterparts (Alsharawy et al., 2021). The present study observed a similar phenomenon. Additionally, nurses with diplomas rated the risk of falling significantly higher than their degree-holding counterparts. This contrasts with findings from other studies, which suggest that individuals with higher educational levels tend to perceive higher probabilities of risk (Pförtner and Hower, 2022, Rattay et al., 2021, Röpcke et al., 2024). It is not certain why this difference occurred. One possible explanation is that nurses with diplomas may have received training that emphasised patient safety and risk during their pre-licensure education.

The final source of judgement variation is cognitive bias. Availability bias was observed in 42.39% of nurses, who rated scenarios approximately 3% lower than their counterparts. Although this difference was small, the findings align with previous research showing that cognitive bias affects nurses’ decision-making and practice (Jala et al., 2023, Whyte et al., 2022). Availability bias increases with the ease of recalling relevant examples (Tversky and Kahneman, 1973). Therefore, if nurses recalled recent instances where patients with similar characteristics fell, their fall risk estimates would increase, while recalling instances where such patients did not fall would lead to underestimation. In this study, nurses probably recalled situations where patients with similar characteristics did not fall, which contributed to their underestimation of risk.

Although base-rate neglect and belief bias did not appear to impact nurses’ judgements, attention should be given to their susceptibility to these cognitive biases. Our findings revealed that 93.44% of nurses exhibited base-rate neglect, while 69.25% were affected by belief bias. These findings are consistent with those of Al-Moteri et al. (Al-Moteri et al., 2020), who reported that 63% of their sample showed cognitive bias. Base-rate neglect occurs when nurses disregard underlying probabilities, while belief bias arises when nurses base their judgement on the believability of conclusions rather than logic. Given that nurses often express discomfort with probabilities (Lavoie et al., 2020), and rely on intuition (Honda et al., 2015), these findings are not surprising. Correct application of probability theory and logical reasoning is essential for accurate judgement and forecasting (Wright et al., 2009). Therefore, these cognitive biases could still affect nurses’ judgements and interventions. Future studies using more reliable instruments would provide a clearer understanding of the impact of these biases.

## Implications for nursing management and research

The results of the regression tree analysis suggest that nurses do not assign equal weight to each risk factor. A comparison of nurses’ judgements with existing fall assessment scales highlights clear distinctions. For example, in the Morse Fall Scale (Morse et al., 1989), patients’ mental status (e.g., forgetting limitations and moving independently) is assigned only half the points (15 points) compared to the use of ambulatory aids (30 points). However, nurses in this study regarded the former as a greater risk. Similarly, in the STRATIFY Risk Assessment Tool (Oliver et al., 1997), all risk factors are equally weighted, yet nurses did not treat them as such, with the weight assigned varying by clinical specialty. Nurses’ contextual knowledge, which has not been integrated into existing models, may provide valuable insights for improving fall assessment scales (Önkal-Atay et al., 2004).

At the same time, nurses and healthcare organisations may need to reconsider their approach to fall prevention. A goal of zero falls may be unrealistic, particularly in rehabilitation units where patients push their physical and cognitive limits to regain optimal functioning (Singh et al., 2019). In their efforts to prevent falls, nurses often instruct patients to call for assistance when ambulating. However, this practice may restrict patients’ independence and dignity (Kerr et al., 2023). A shift in focus toward preventing recurrent falls and minimising fall-related injuries may be more practical and patient-centred.

Finally, this study emphasises the importance of reducing nurses’ cognitive biases. To achieve this, organisations should offer training in probabilistic reasoning and logical inference to enhance nurses’ reasoning strategies (Čavojová et al., 2020). Such training can help reduce base-rate neglect, belief bias, and availability bias. Additionally, encouraging slow thinking and fostering judgement verification through peer consultation can further minimise cognitive bias. Organisations should also establish a framework that supports a reflective clinical culture.

## Limitations

The current study has several limitations. First, the scenarios were based on a limited set of fall risks, which may not fully capture the complexity of nurses’ judgement strategies. Second, only three types of cognitive bias were examined, and there are many other types that may influence nurses’ judgements. Further research is needed to explore the impact of additional cognitive biases and identify their sources of variation. Third, the reliability of the cognitive bias scales was low, partly due to small variances in nurses’ responses, which tended to concentrate around specific answers. The low reliability of these scales has been noted in other studies, with potential remedial solutions suggested by Berthet (2021). Developing more reliable, context-specific scales is essential for improving the accuracy of cognitive bias measurement.

## Conclusions

Judging fall risk is a complex process that involves identifying risk factors and assessing their relative importance, while also considering potential synergies and counteractions between them. Contextual factors further influence this judgement, as the significance of risk factors can vary across different clinical areas. Due to this complexity, human judgement is essential, although it is this very judgement that may contribute to clinical variation. While variation in clinical judgement is generally seen as undesirable because it can lead to inconsistent practices, not all variation results in unjustified care. For example, variation in nurses’ fall risk judgements may be justified when it reflects personalised, context-specific nursing care. In contrast, errors arising from cognitive bias, which leads to inconsistent judgements, are unjustified. Nurses’ clinical judgement is directly linked to the effectiveness of fall prevention measures. Consequently, healthcare organisations should provide targeted training to enhance nurses’ contextual expertise while mitigating cognitive biases.

## Data availability

Data not available

The participants were assured raw data would remain confidential and would not be shared.

## Acknowledgements

The authors would like to express their gratitude to Dr. Marjolein Fokkema for her assistance in conducting a linear mixed-effects model tree and a 10-fold cross-validation.

## Supplementary Materials

The supplementary file includes patient scenarios used in the study, factor analysis results of the cognitive scales and reliability coefficients, response frequencies for cognitive biases, and Spearman correlation coefficients between variables.

## References

Agency for Healthcare Research and Quality, 2013. Preventing falls in hospitals: A toolkit for improving quality of care.

Ainuddin, H.A., Romli, M.H., Salim, M.S.F., Hamid, T.A.T.A., Mackenzie, L., 2019. Stroke rehabilitation for falls and Risk of Falls in Southeast Asia: A Scoping Review with Stakeholders’ Consultation. Age and Ageing 48 (Supplement_4), iv4–iv5, 10.1093/ageing/afz164.16.

Al-Moteri, M., Cooper, S., Symmons, M., Plummer, V., 2020. Nurses’ cognitive and perceptual bias in the identification of clinical deterioration cues. Australian Critical Care 33 (4), 333–342, 10.1016/j.aucc.2019.08.006.

Al-Moteri, M., Plummer, V., Cooper, S., 2022. Decision-Making Errors During Recognizing and Responding to Clinical Deterioration: Gaze Path-Cued Retrospective Think-Aloud. Clinical Simulation in Nursing 73, 29–36, 10.1016/j.ecns.2022.08.002.

Alsharawy, A., Spoon, R., Smith, A., Ball, S., 2021. Gender Differences in Fear and Risk Perception During the COVID-19 Pandemic. Front Psychol 12, 689467, 10.3389/fpsyg.2021.689467.

Althnian, A., AlSaeed, D., Al-Baity, H., Samha, A., Dris, A., Alzakari, N., Abou Elwafa, A., Kurdi, H., 2021. Impact of dataset size on classification performance: An empirical evaluation in the medical domain. Applied Sciences 11, 796, 10.3390/app11020796.

Alvarado, N., McVey, L., Wright, J., Healey, F., Dowding, D., Cheong, V.L., Gardner, P., Hardiker, N., Lynch, A., Zaman, H., Smith, H., Randell, R., 2023. Exploring variation in implementation of multifactorial falls risk assessment and tailored interventions: a realist review. BMC Geriatrics 23 (1), 381, 10.1186/s12877-023-04045-3.

Berthet, V., 2021. The Measurement of Individual Differences in Cognitive Biases: A Review and Improvement. Frontiers in Psychology 12, 10.3389/fpsyg.2021.630177.

Blanco, F., 2020. Cognitive bias. In: Vonk, J., Todd., S. (Eds.), Encyclopedia of animal cognition and behavior. Cham.

Brown, G.D., Largey, A., McMullan, C., 2021. The impact of gender on risk perception: Implications for EU member states’ national risk assessment processes. International Journal of Disaster Risk Reduction 63, 102452, 10.1016/j.ijdrr.2021.102452.

Čavojová, V., Šrol, J., Jurkovič, M., 2020. Why should we try to think like scientists? Scientific reasoning and susceptibility to epistemically suspect beliefs and cognitive biases. Applied Cognitive Psychology 34 (1), 85–95, 10.1002/acp.3595.

Ceschi, A., Costantini, A., Sartori, R., Weller, J., Di Fabio, A., 2019. Dimensions of decision-making: An evidence-based classification of heuristics and biases. Personality and Individual Differences 146, 188–200, 10.1016/j.paid.2018.07.033.

Choi, J., Lee, S., Park, E., Ku, S., Kim, S., Yu, W., Jeong, E., Park, S., Park, Y., Kim, S.R., 2024. Congruency and its related factors between patients’ fall risk perception and nurses’ fall risk assessment in acute care hospitals. Journal of Nursing Scholarship 56 (4), 507–516, 10.1111/jnu.12964.

Connor, J., Flenady, T., Massey, D., Dwyer, T., 2023. Clinical judgement in nursing – An evolutionary concept analysis. Journal of Clinical Nursing 32 (13-14), 3328–3340, 10.1111/jocn.16469.

Croskerry, P., 2003. The importance of cognitive errors in diagnosis and strategies to minimize them. Acad Med 78 (8), 775–780, 10.1097/00001888-200308000-00003.

Croskerry, P., Ryle, C.A., 2019. Croskerry’s List of 50 Common Biases: 50 Cognitive and Affective Biases in Medicine (Alphabetically). In, Risk and Reasoning in Clinical Diagnosis. Oxford University Press, pp. 0.

Crowley, R.S., Legowski, E., Medvedeva, O., Reitmeyer, K., Tseytlin, E., Castine, M., Jukic, D., Mello-Thoms, C., 2013. Automated detection of heuristics and biases among pathologists in a computer-based system. Advances in Health Sciences Education 18 (3), 343–363, 10.1007/s10459-012-9374-z.

Deandrea, S., Lucenteforte, E., Bravi, F., Foschi, R., La Vecchia, C., Negri, E., 2010. Risk factors for falls in community-dwelling older people: A systematic review and meta-analysis. Epidemiology 21 (5), 658–668,

Erceg, N., Galic, Z., Bubić, A., 2022. Normative responding on cognitive bias tasks: Some evidence for a weak rationality factor that is mostly explained by numeracy and actively open-minded thinking. Intelligence 90, 101619, 10.1016/j.intell.2021.101619.

Essa, C.D., Victor, G., Khan, S.F., Ally, H., Khan, A.S., 2023. Cognitive biases regarding utilization of emergency severity index among emergency nurses. The American Journal of Emergency Medicine 73, 63–68, 10.1016/j.ajem.2023.08.021.

Fernández-de-Maya, J., Richart-Martínez, M., 2012. Variability of clinical practice in nursing: An integrative review. ACTA Paulista de Enfermagem 25 (5), 809–816,

Ferrario, C.G., 2003. Experienced and less-experienced nurses’ diagnostic reasoning: Implications for fostering students’ critical thinking. International Journal of Nursing Terminologies and Classifications 14 (2), 41–52, 10.1111/j.1744-618X.2003.tb00059.x.

Fhon, J.R.S., Silva, A.R.F., Lima, E.F.C., Santos Neto, A.P.D., Henao-Castaño Á, M., Fajardo-Ramos, E., Püschel, V.A.A., 2023. Association between sarcopenia, falls, and cognitive impairment in older people: A systematic review with meta-analysis. International Journal of Environmental Research and Public Health 20 (5), 4156, 10.3390/ijerph20054156.

Fokkema, M., Smits, N., Zeileis, A., Hothorn, T., Kelderman, H., 2018. Detecting treatment-subgroup interactions in clustered data with generalized linear mixed-effects model trees. Behav Res Methods 50 (5), 2016–2034, 10.3758/s13428-017-0971-x.

Francis-Coad, J., Hill, A.-M., Jacques, A., Chandler, A.M., Richey, P.A., Mion, L.C., Shorr, R.I., 2020. Association between characteristics of Iinjurious falls and fall preventive interventions in acute medical and surgical units. The Journals of Gerontology: Series A 75 (10), e152–e158, 10.1093/gerona/glaa032.

Grömping, U., 2018. R Package DoE.base for factorial experiments. Journal of Statistical Software 85 (5), 1–41, 10.18637/jss.v085.i05.

Hancock, H.C., Mason, J.M., Murphy, J.J., 2012. Using the method of judgement analysis to address variations in diagnostic decision making. BMC Res Notes 5, 139, 10.1186/17560500-5-139.

Harris, C.R., Jenkins, M., 2006. Gender Differences in Risk Assessment: Why do Women Take Fewer Risksthan Men? Judgment and Decision Making 1 (1), 48–63, 10.1017/S1930297500000346.

Heinzmann, J., Rossen, M.L., Efthimiou, O., Baumgartner, C., Wertli, M.M., Rodondi, N., Aubert, C.E., Liechti, F.D., 2024. Risk factors for falls among hospitalized medical patients – A systematic review and meta-analysis. Archives of Physical Medicine and Rehabilitation, 10.1016/j.apmr.2024.06.015.

Honda, H., Ogawa, M., Murakoshi, T., Masuda, T., Utsumi, K., Nei, D., Wada, Y., 2015. Variation in risk judgment on radiation contamination of food: Thinking trait and profession. Food Quality and Preference 46, 119–125, 10.1016/j.foodqual.2015.07.014.

Hong, S., Kim, J.S., Choi, Y.A., 2024. Predictive Validity of the Johns Hopkins Fall Risk Assessment Tool for Older Patients in Stroke Rehabilitation. Healthcare (Basel) 12 (7), 10.3390/healthcare12070791.

Innab, A.M., 2022. Nurses’ perceptions of fall risk factors and fall prevention strategies in acute care settings in Saudi Arabia. Nursing Open 9 (2), 1362–1369, 10.1002/nop2.1182.

Inoue, S., Otaka, Y., Mori, N., Matsuura, D., Tsujikawa, M., Kawakami, M., Kondo, K., 2024. Blind spots in hospital fall prevention: Falls in stroke patients occurred not only in those at a high risk of falling. Journal of the American Medical Directors Association 25 (1), 160–166.e161, 10.1016/j.jamda.2023.10.034.

Jala, S., Fry, M., Elliott, R., 2023. Cognitive bias during clinical decision-making and its influence on patient outcomes in the emergency department: A scoping review. Journal of Clinical Nursing 32 (19-20), 7076–7085, 10.1111/jocn.16845.

Japan Hospital Association, 2023. QI project report. Tokyo.

Kahneman, D., Tversky, A., 1982. On the psychology of prediction. In: Kahneman, D., Slovic, P., Tversky, A. (Eds.), Judgment under Uncertainty: Heuristics and Biases. Cambridge University Press, Cambridge, pp. 48–68.

Kato, Y., Kitamura, S., Katoh, M., Hirano, A., Senjyu, Y., Ogawa, M., Maeda, H., Mukaino, M., Hirano, S., Sakurai, H., Shibata, S., Otaka, Y., 2022. Stroke patients with nearly independent transfer ability are at high risk of falling. J Stroke Cerebrovasc Dis 31 (1), 106169, 10.1016/j.jstrokecerebrovasdis.2021.106169.

Kerr, L., Newman, P., Russo, P., 2023. ’I don’t want to impose on anybody’: Older people and their families discuss their perceptions of risk, cause and care in the context of falls. Int J Older People Nurs 18 (6), e12578, 10.1111/opn.12578.

Killip, S., Mahfoud, Z., Pearce, K., 2004. What is an intracluster correlation coefficient? Crucial concepts for primary care researchers. Ann Fam Med 2 (3), 204–208, 10.1370/afm.141.

Kim, Y., Park, I., Kang, S., 2018. Age and gender differences in health risk perception. Cent Eur J Public Health 26 (1), 54–59, 10.21101/cejph.a4920.

Kobayashi, K., Imagama, S., Ando, K., Inagaki, Y., Suzuki, Y., Nishida, Y., Nagao, Y., Ishiguro, N., 2017. Analysis of falls that caused serious events in hospitalized patients. Geriatrics and Gerontology International 17 (12), 2403–2406, 10.1111/ggi.13085.

Korteling, J.E., Paradies, G., Sassen-van Meer, J., 2023. Cognitive bias and how to improve sustainable decision making. Frontiers in Psychology 14, 1129835, 10.3389/fpsyg.2023.1129835.

Korteling, J.E., Toet, A., 2021. Cognitive biases. In: Della Sala, S. (Ed.), Encyclopedia of behavioural neuroscience. Elsevier, pp. 610–619.

Kumle, L., Võ, M.L.-H., Draschkow, D., 2021. Estimating power in (generalized) linear mixed models: An open introduction and tutorial in R. Behavior Research Methods 53, 2528–2543,

Lavoie, P., Clarke, S.P., Clausen, C., Purden, M., Emed, J., Mailhot, T., Fontaine, G., Frunchak, V., 2020. Nurses’ judgments of patient risk of deterioration at change-of-shift handoff: Agreement between nurses and comparison with early warning scores. Heart & Lung 49 (4), 420–425, 10.1016/j.hrtlng.2020.02.037.

Lee, F.S., Sararaks, S., Yau, W.K., Ang, Z.Y., Jailani, A.S., Abd Karim, Z., Naing, L., Krishnan, T., Chu, A.R., Junus, S., Ahmad, M.S., Sapiee, N., Veloo, V.W., Manoharan, S., A. Hamid, M., 2022. Fall determinants in hospitalised older patients: a nested case control design - incidence, extrinsic and intrinsic risk in Malaysia. BMC Geriatrics 22 (1), 179, 10.1186/s12877-022-02846-6.

Martin, K., Bickle, K., Lok, J., 2022. Investigating the impact of cognitive bias in nursing documentation on decision-making and judgement. International Journal of Mental Health Nursing 31 (4), 897–907, 10.1111/inm.12997.

Martin, K.L., Blizzard, L., Srikanth, V.K., Wood, A., Thomson, R., Sanders, L.M., Callisaya, M.L., 2013. Cognitive function modifies the effect of physiological function on the risk of multiple falls--a population-based study. J Gerontol A Biol Sci Med Sci 68 (9), 1091–1097, 10.1093/gerona/glt010.

McDonnell, T., Nicholson, E., Bury, G., Collins, C., Conlon, C., De Brún, A., Doherty, E., McAuliffe, E., 2023. The role of contextual factors in decision-making by General Practitioners on paediatric referral to the Emergency Department in Ireland: A Discrete Choice Experiment. Health Policy 132, 104813, 10.1016/j.healthpol.2023.104813.

Milisen, K., Coussement, J., Flamaing, J., Vlaeyen, E., Schwendimann, R., Dejaeger, E., Surmont, K., Boonen, S., 2012. Fall prediction cccording to nurses’ clinical judgment: Differences between medical, surgical, and geriatric wards. Journal of the American Geriatrics Society 60 (6), 1115–1121, 10.1111/j.1532-5415.2012.03957.x.

Moon, S., Chung, H.S., Kim, Y.J., Kim, S.J., Kwon, O., Lee, Y.G., Yu, J.M., Cho, S.T., 2021. The impact of urinary incontinence on falls: A systematic review and meta-analysis. PLoS One 16 (5), e0251711, 10.1371/journal.pone.0251711.

Morse, J.M., Morse, R.M., Tylko, S.J., 1989. Development of a scale to identify the fall-prone patient. Canadian Journal on Aging / La Revue canadienne du vieillissement 8 (4), 366–377, 10.1017/S0714980800008576.

Nakanishi, T., Ikeda, T., Nakamura, T., Yamanouchi, Y., Chikamoto, A., Usuku, K., 2021. Development of an algorithm for assessing fall risk in a Japanese inpatient population. Scientific Reports 11 (1), 17993, 10.1038/s41598-021-97483-1.

Noguchi, N., Chan, L., Cumming, R.G., Blyth, F.M., Naganathan, V., 2016. A systematic review of the association between lower urinary tract symptoms and falls, injuries, and fractures in community-dwelling older men. Aging Male 19 (3), 168–174, 10.3109/13685538.2016.1169399.

O’Sullivan, E.D., Schofield, S.J., 2018. Cognitive bias in clinical medicine. J R Coll Physicians Edinb 48 (3), 225–232, 10.4997/jrcpe.2018.306.

Oliver, D., Britton, M., Seed, P., Martin, F.C., Hopper, A.H., 1997. Development and evaluation of evidence based risk assessment tool (STRATIFY) to predict which elderly inpatients will fall: case-control and cohort studies. BMJ 315 (7115), 1049–1053, 10.1136/bmj.315.7115.1049.

Önkal-Atay, D., Thomson, M.E., Pollock, A.C., 2004. Judgmental forecasting. In: Michael P. Clements, David F. Hendry (Eds.), A Companion to Economic Forecasting. Wiley-Blackwell, New Jersey, US, pp. 133–151.

Oren, G., Jolkovsky, S., Tal, S., 2022. Falls in oldest-old adults hospitalized in acute geriatric ward. Eur Geriatr Med 13 (4), 859–866, 10.1007/s41999-022-00660-2.

Perell, K.L., Nelson, A., Goldman, R.L., Luther, S.L., Prieto-Lewis, N., Rubenstein, L.Z., 2001. Fall risk assessment measures: An analytic review. The Journals of Gerontology: Series A 56 (12), M761–M766, 10.1093/gerona/56.12.M761.

Pförtner, T.K., Hower, K.I., 2022. Educational inequalities in risk perception, perceived effectiveness, trust and preventive behaviour in the onset of the COVID-19 pandemic in Germany. Public Health 206, 83–86, 10.1016/j.puhe.2022.02.021.

Prabhakaran, K., Gogna, S., Pee, S., Samson, D.J., Con, J., Latifi, R., 2020. Falling again? Falls in geriatric adults-risk factors and outcomes associated with recidivism. J Surg Res 247, 66–76, 10.1016/j.jss.2019.10.041.

Rajput, D., Wang, W.-J., Chen, C.-C., 2023. Evaluation of a decided sample size in machine learning applications. BMC Bioinformatics 24 (1), 48, 10.1186/s12859-023-05156-9.

Rattay, P., Michalski, N., Domanska, O.M., Kaltwasser, A., De Bock, F., Wieler, L.H., Jordan, S., 2021. Differences in risk perception, knowledge and protective behaviour regarding COVID-19 by education level among women and men in Germany. Results from the COVID-19 Snapshot Monitoring (COSMO) study. PLoS One 16 (5), e0251694, 10.1371/journal.pone.0251694.

Rice, H., Garabedian, P.M., Shear, K., Bjarnadottir, R.I., Burns, Z., Latham, N.K., Schentrup, D., Lucero, R.J., Dykes, P.C., 2022. Clinical decision support for fall prevention: defining end-user needs. Appl Clin Inform 13 (3), 647–655, 10.1055/s-0042-1750360.

Röpcke, A., Brinkmann, C., Neumann-Böhme, S., Sabat, I., Barros, P.P., Schreyögg, J., Torbica, A., Brouwer, W., Hajek, A., Stargardt, T., 2024. Understanding health risk perception: insights from an eight-country panel study during the COVID-19 pandemic. Journal of Public Health, 10.1007/s10389-024-02351-7.

Sakai, M., Rossaneis, A., Ângela, M., Haddad, F.L., do Carmo, M., Dagmar, W.V., 2016. Risk of bed falls in Adult patients and prevention measures. Journal of Nursing UFPE / Revista de Enfermagem UFPE 10, 4720–4726, 10.5205/reuol.8200-71830-3-SM.1006sup201602.

Saposnik, G., Redelmeier, D., Ruff, C.C., Tobler, P.N., 2016. Cognitive biases associated with medical decisions: a systematic review. BMC Medical Informatics and Decision Making 16 (1), 138, 10.1186/s12911-016-0377-1.

Satoh, M., Miura, T., Shimada, T., 2023. Development and evaluation of a simple predictive model for falls in acute care setting. J Clin Nurs 32 (17-18), 6474–6484, 10.1111/jocn.16680.

Satoh, M., Miura, T., Shimada, T., Hamazaki, T., 2022. Risk stratification for early and late falls in acute care settings. Journal of Clinical Nursing, 10.1111/jocn.16267.

Shao, L., Shi, Y., Xie, X.-Y., Wang, Z., Wang, Z.-A., Zhang, J.-E., 2023. Incidence and Risk Factors of Falls Among Older People in Nursing Homes: Systematic Review and Meta-Analysis. Journal of the American Medical Directors Association 24 (11), 1708–1717, 10.1016/j.jamda.2023.06.002.

Shinnick, M.A., Cabrera-Mino, C., 2021. Predictors of nursing clinical judgment in simulation. Nurs Educ Perspect 42 (2), 107–109, 10.1097/01.Nep.0000000000000604.

Singh, H., Craven, B.C., Flett, H.M., Kerry, C., Jaglal, S.B., Silver, M.P., Musselman, K.E., 2019. Factors influencing fall prevention for patients with spinal cord injury from the perspectives of administrators in Canadian rehabilitation hospitals. BMC Health Serv Res 19 (1), 391, 10.1186/s12913-019-4233-8.

Sordo, M., Zeng, Q., 2005. On sample size and classification accuracy: A performance comparison. In: Biological and Medical Data Analysis. Springer Berlin Heidelberg, Berlin, Heidelberg, pp. 193–201.

Stamp, K.D., 2012. How nurse practitioners make decisions regarding coronary heart disease risk: a social judgment analysis. Int J Nurs Knowl 23 (1), 29–40, 10.1111/j.2047-3095.2011.01196.x.

Sutherland, K., Levesque, J.F., 2020. Unwarranted clinical variation in health care: Definitions and proposal of an analytic framework. J Eval Clin Pract 26 (3), 687–696, 10.1111/jep.13181.

Taggart, S., Skylas, K., Brannelly, A., Fairbrother, G., Knapp, M., Gullick, J., 2021. Using a clinical judgement model to understand the impact of validated pain assessment tools for burn clinicians and adult patients in the ICU: A multi-methods study. Burns 47 (1), 110–126, 10.1016/j.burns.2020.05.032.

Takase, M., 2023. Falls as the result of interplay between nurses, patient and the environment: Using text-mining to uncover how and why falls happen. International Journal of Nursing Sciences 10 (1), 30–37, 10.1016/j.ijnss.2022.12.003.

Takase, M., Kisanuki, N., Nakayoshi, Y., Uemura, C., Sato, Y., Yamamoto, M., 2024. Exploring nurses’ clinical judgment concerning the relative importance of fall risk factors: A mixed method approach using the Q Methodology. International Journal of Nursing Studies 153, 104720, 10.1016/j.ijnurstu.2024.104720.

Tanner, C.A., 2006. Thinking like a nurse: a research-based model of clinical judgment in nursing. The Journal of nursing education 45 6, 204–211,

Thompson, C., Bucknall, T., Estabrookes, C.A., Hutchinson, A., Fraser, K., de Vos, R., Binnecade, J., Barrat, G., Saunders, J., 2009. Nurses’ critical event risk assessments: a judgement analysis. J Clin Nurs 18 (4), 601–612, 10.1111/j.1365-2702.2007.02191.x.

Tversky, A., Kahneman, D., 1973. Availability: A heuristic for judging frequency and probability. Cognitive Psychology 5 (2), 207–232, 10.1016/0010-0285(73)90033-9.

Tzeng, H.M., Yin, C.Y., 2013. Frequently observed risk factors for fall-related injuries and effective preventive interventions: a multihospital survey of nurses’ perceptions. Journal of Nursing Care Quality 28 (2), 130–138, 10.1097/NCQ.0b013e3182780037.

Vlaeyen, E., Poels, J., Colemonts, U., Peeters, L., Leysens, G., Delbaere, K., Dejaeger, E., Dobbels, F., Milisen, K., 2021. Predicting falls in nursing homes: A prospective multicenter cohort study comparing fall history, staff clinical judgment, the care home falls screen, and the fall risk classification algorithm. Journal of the American Medical Directors Association 22 (2), 380–387, 10.1016/j.jamda.2020.06.037.

Wabe, N., Meulenbroeks, I., Huang, G., Silva, S.M., Gray, L.C., Close, J.C.T., Lord, S., Westbrook, J.I., 2024. Development and internal validation of a dynamic fall risk prediction and monitoring tool in aged care using routinely collected electronic health data: a landmarking approach. Journal of the American Medical Informatics Association 31 (5), 1113–1125, 10.1093/jamia/ocae058.

Whyte, S., Rego, J., Fai Chan, H., Chan, R.J., Yates, P., Dulleck, U., 2022. Cognitive and behavioural bias in advance care planning. Palliat Care Soc Pract 16, 26323524221092458, 10.1177/26323524221092458.

World Health Organization, 2007. WHO Global Report on Falls Prevention in Older Age. Accessed: April, 2022. https://extranet.who.int/agefriendlyworld/wp-content/uploads/2014/06/WHo-Global-report-on-falls-prevention-in-older-age.pdf.

Wright, G., Bolger, F., Rowe, G., 2009. Expert judgement of probability and risk. In: Williams, T.M., Samset, K., Sunnevåg, K.J. (Eds.), Making essential choices with scant information: Front-end decision making in major projects. Palgrave Macmillan UK, London, pp. 213–229.

Wright, G., Rowe, G., Bolger, F., Gammack, J., 1994. Coherence, calibration, and expertise in judgmental probability forecasting. Organizational Behavior and Human Decision Processes 57 (1), 1–25, 10.1006/obhd.1994.1001.

Wu, M.W., Lee, T.T., Lai, S.M., Huang, C.Y., Chang, T.H., 2019. Evaluation of electronic health records on the nursing process and patient outcomes regarding fall and pressure injuries. Comput Inform Nurs 37 (11), 573–582, 10.1097/cin.0000000000000548.

Xu, Q., Ou, X., Li, J., 2022. The risk of falls among the aging population: A systematic review and meta-analysis. Front Public Health 10, 902599, 10.3389/fpubh.2022.902599.

Yang, H., Thompson, C., 2016. Capturing judgement strategies in risk assessments with improved quality of clinical information: How nurses’ strategies differ from the ecological model. BMC Medical Informatics and Decision Making 16 (1), 7, 10.1186/s12911-016-0243-1.

Yasan, C., Burton, T., Tracey, M., 2020. Nurses’ documentation of falls prevention in a patient centred care plan in a medical ward. Australian Journal of Advanced Nursing 37 (2), 19–24, 10.37464/2020.372.103.

