## Supplementary tables 1-4 for "Nurses’ Fall Risk Judgements - Cognitive Biases and Contextual Factors Explaining Variability: A multi-centre cross-sectional study" (from another version of this preprint: version 1, 2024-12-05)

Supplementary Table 1. Patient scenarios used in the study

| Scenario No. | Mobility issues | Cognitive ability and personality (Does not call nurses) | Urinary issues | Presence of tubes or drains | Medication use | Age |
|---|---|---|---|---|---|---|
| 1 | Need assistance in transfer | No cognitive impairment, but reserved and does not call nurses when necessary | Nocturia | No tube or drain inserted | Sleeping pill | Less than 65 years old |
| 2 | Independent | No cognitive impairment and calls nurses when necessary | Nocturia | One tube or drain inserted | Sleeping pill | 80 years old and over |
| 3 | Walks alone but unstable | No cognitive impairment, but reserved and does not call nurses when necessary | Nocturia | One tube or drain inserted | Anti-hypertensive | 65-79 years old |
| 4 | Walks alone but unstable | Impaired cognition and does not call nurses when necessary | No urinary problems | One tube or drain inserted | Sleeping pill | Less than 65 years old |
| 5 | Walks alone but unstable | No cognitive impairment and calls nurses when necessary | No urinary problems | Multiple tubes and/or drains inserted | Anti-hypertensive | 80 years old and over |
| 6 | Needs assistance in transfer | No cognitive impairment and calls nurses when necessary | Frequency | One tube or drain inserted | Anti-hypertensive | Less than 65 years old |
| 7 | Walk alone but unstable | Impaired cognition and does not call nurses when necessary | Nocturia | No tube or drain inserted | Antibiotic | 80 years old and over |
| 8 | Needs assistance in transfer | Impaired cognition and does not call nurses when necessary | Frequency | Multiple tubes and/or drains inserted | Sleeping pill | 80 years old and over |
| 9 | Independent | No cognitive impairment, but reserved and does not call nurses | Frequency | No tube or drain inserted | Anti-hypertensive | 80 years old and over |

| | | | | | | |
|---|---|---|---|---|---|---|
| | | when necessary | | | | |
| 10 | Walks alone but unstable | No cognitive impairment, but reserved and does not call nurses when necessary | Frequency | Multiple tubes and/or drains inserted | Antibiotic | Less than 65 years old |
| 11 | Walks alone but unstable | No cognitive impairment and calls nurses when necessary | Frequency | No tube or drain inserted | Sleeping pill | 65-79 years old |
| 12 | Needs assistance in transfer | No cognitive impairment and calls nurses when necessary | Nocturia | Multiple tubes and/or drains inserted | Antibiotic | 65-79 years old |
| 13 | Independent | No cognitive impairment, but reserved and does not call nurses when necessary | No urinary problems | Multiple tubes and/or drains inserted | Sleeping pill | 65-79 years old |
| 14 | Independent | Impaired cognition and does not call nurses when necessary | Frequency | One tube or drain inserted | Antibiotic | 65-79 years old |
| 15 | Needs assistance in transfer | No cognitive impairment, but reserved and does not call nurses when necessary | No urinary problems | One tube or drain inserted | Antibiotic | 80 years old and over |
| 16 | Needs assistance in transfer | Impaired cognition and does not call nurses when necessary | No urinary problems | No tube or drain inserted | Anti-hypertensive | 65-79 years old |
| 17 | Needs assistance in transfer | No cognitive impairment and calls nurses when necessary | No urinary problems | No tube or drain inserted | Antibiotic | Less than 65 years old |
| 18 | Needs assistance in transfer | Impaired cognition and does not call nurses when necessary | Nocturia | Multiple tubes and/or drains inserted | Anti-hypertensive | Less than 65 years old |

Supplementary Table 2. Factor analysis of the cognitive scales and reliability coefficients

| Variables | Factor 1 | Factor 2 | Factor 3 |
|---|---|---|---|
| BN item1 | 0.665 | | |
| BN item2 | 0.740 | | |
| BN item3 | 0.593 | | |
| AB item1 | | 0.670 | |
| AB item2 | | 0.659 | |
| AB item3 | | 0.622 | |
| BB item1 | | | 0.651 |
| BB item2 | | | 0.697 |
| BB item3 | | | 0.436 |
| Eigenvalue | 2.169 | 2.030 | 1.587 |
| Variance explained (Cumulative variance) | 0.241 | 0.226 (0.467) | 0.176 (0.643) |
| KR 20 coefficients | 0.323 | 0.562 | 0.434 |

Note: BN: Base-rate neglect, AB: Availability bias, BB: Belief bias
Bartlett's test of sphericity: Chi-squared (36) = 804.64, p < .001, The Kaiser, Meyer, Olkin (KMO) = 0.59.

Supplementary Table 3. Frequency of the responses in cognitive bias

| Total score | Base-rate neglect | Belief bias | Availability bias |
|---|---|---|---|
| 0 | 4 (1.19%) | 28 (8.36%) | 98 (29.25%) |
| 1 | 16 (4.78%) | 68 (20.30%) | 93 (27.76%) |
| 2 | 83 (24.78%) | 113 (33.73%) | 88 (26.27%) |
| 3 | 230 (68.66%) | 119 (35.52%) | 54 (16.12%) |
| Missing | 2 (0.60%) | 7 (2.09%) | 2 (0.60%) |
| Mean (SD) | 2.62 (0.64) | 1.99 (0.95) | 1.29 (1.06) |

Note: Means and the standard deviations were calculated after replacing missing values with the median values.

Supplementary Table 4. Spearman correlation coefficients between variables

| | Experience | Age | Education | Sex | Base-rate neglect | Belief bias | Availability bias |
|---|---|---|---|---|---|---|---|
| Age | 0.921* | | | | | | |
| Education | -0.291* | -0.271* | | | | | |
| Sex | -0.004 | 0.028 | 0.073 | | | | |
| Base-rate neglect | -0.042 | -0.074 | -0.142* | -0.090 | | | |
| Belief bias | 0.100 | 0.061 | -0.060 | 0.025 | 0.056 | | |
| Availability bias | 0.098 | 0.113* | 0.025 | 0.105 | -0.050 | 0.072 | |
| Ns Judgement (Mean) | -0.052 | -0.079 | -0.138* | -0.150* | 0.099 | -0.029 | -0.166* |

Note: * p<0.05. Missing values were replaced with the median values in the continuous/ordinal variables, and the most representative category in the nominal variables.
The dummy variables are as follows: Education (0: Diploma, 1: Degree), Sex (0: Women, 1: Men). The scores of the bias scales ranged from 0 to 3.
